# Genetic Insights into the Microevolutionary Dynamics and Early Introductions of Human Monkeypox Virus in Mexico

**DOI:** 10.1101/2023.08.26.23294674

**Authors:** Israel Gómez-Sánchez, Hugo G. Castelán-Sánchez, León P. Martínez-Castilla, Juan Manuel Hurtado-Ramírez, Gamaliel López-Leal

**Affiliations:** Laboratorio de Biología Computacional y Virómica Integrativa, Centro de Investigación en Dinámica Celular, Universidad Autónoma del Estado de Morelos, Cuernavaca, Morelos, México; Grupo de Genómica y Dinámica Evolutiva de Microorganismos Emergentes. Programa de Investigadoras e Investigadores por México. Consejo Nacional de Ciencia y Tecnología. Ciudad de México, México; Instituto de Biotecnología, Universidad Nacional Autónoma de México, Cuernavaca, Morelos, México

**Keywords:** Viruses, Monkeypox, Natural Selection, Virus Evolution

## Abstract

The recent global outbreak of Monkeypox, caused by the Monkeypox virus (MPVX) emerged in Europe in 2022 and rapidly spread to over 40 countries. The Americas are currently facing the highest impact, reporting over 50,000 cases by early 2023. Here, we analyze 880 MPXV isolates worldwide to gain insights into the evolutionary patterns and initial introduction events of the virus in Mexico. We found that MPXV entered Mexico on multiple occasions, from the United Kingdom, Portugal, and Canada, and subsequently spread locally in different regions of Mexico. Additionally, we show that MPXV has an open pangenome, highlighting the role of gene turnover in shaping its genomic diversity, rather than SNPs variations, which do not contribute significantly to genome diversity. Although the genome presents multiple SNP sites in coding regions, these remain under purifying selection, suggesting their evolutionary conservation. One notable exception is the amino acid site 63 of the protein encoded by the Cop-A4L gene, intricately related to viral maturity, for which we picked up a strong signal of positive selection. Ancestral state reconstruction deduced that the ancestral state at site 63 corresponds to the amino acid valine, present only in isolates of clade I. However, the isolates of the current outbreak evolved for threonine at site 63. Finally, our findings contribute to the knowledge of the evolutionary processes of the Monkeypox virus.

## INTRODUCTION

Monkeypox virus (MPXV) is a double-stranded DNA virus that belongs to the *Poxviridae* family and the *Orthopoxvirus* genus, that causes the infectious Monkeypox viral disease in humans ^1^. MPXV is a close relative of other viruses such as Vaccinia and Cowpox ^2,3^. The first identification of MPXV dates back to 1959 in cynomolgus monkeys ^3^. The first occurrence of monkeypox virus in humans was reported in 1970 in an unvaccinated 9-month-old infant from the Democratic Republic of Congo ^3^. Since then, there have been several major outbreaks in West and Central Africa, including Nigeria, Ghana, and the Central African Republic. Until recently, Monkeypox has been confined to the African continent, with only a handful of sporadic cases in other parts of the world ^4^.

Phylogenetically, MPXV is divided into two main clades. The Central African clade and the West African clade are now designated as clade I and clade II (composed of clades IIa and IIb), respectively. Members of the Central African clade (clade I), also known as the Congo Basin clade, are the most virulent and have been reported mainly in cases in the Democratic Republic of the Congo (DRC) and South Sudan ^5,6^.

On the other hand, cases in Nigeria between 2010 and 2019 were mainly attributed to members of the West African clade ^7^ (Clade IIb). Cases outside Africa, including those currently circulating worldwide, were also all caused by the West African clade. For example, a dramatic increase in MPXV infections outside of Africa was reported in May 2022, resulting in epidemiological outbreaks in different parts of the world ^8^. In less than four months, the number of detected MPXV infections increased to more than 48,000 cases, with 13 deaths ^9^, causing the World Health Organization (WHO) to declare the monkeypox outbreak a global health emergency. By the end of July 2023, 88,600 confirmed cases and 152 deaths had already been reported by the WHO ^9^. However, even though the new outbreak of MPXV is no longer considered a health emergency, countries in the American continent continue to be the most affected by MPXV. At the beginning of 2023 alone, 57,922 confirmed cases were reported in the American continent ^10^. Clearly, this number drastically exceeds the cases reported in Europe and Africa, where 25,699 and 1,195 confirmed cases were reported at the end of 2022, respectively. In Mexico, the first case of MPXV was reported in Mexico City on May 28, 2022. Currently, more than 4,000 confirmed cases have been reported in Mexico. In other words, 131 - 182 cases per month have been reported in just eight months since the first notification, and although the number of new cases has decreased, reporting continued up to late July 2023 ^10^. Furthermore, from the first report of MPXV in Mexico until mid-2023, 30 deaths have been reported. This positions Mexico as the country with the second-highest number of MPXV-related deaths globally ^10^.

The current COVID-19 pandemic has taught us the importance of continuous genomic surveillance of emerging viruses through rapid sequencing and open genome availability. The emergence of the global MPXV outbreak and their rapid propagation worldwide highlights the need for ongoing surveillance and research to better understand the epidemiology, clinical presentation, and pathogenesis of the virus. In this study, we present a comprehensive genomic analysis of MPXV genomes worldwide that sheds light on the evolutionary dynamics and early introduction events of the virus in Mexico. Finally, our findings contribute to the knowledge of the evolutionary processes of the Monkeypox virus.

## MATERIAL AND METHODS

### Collection of genomes

For our study, we downloaded 880 MPXV genomes reported from different parts of the world and publicly available on the GISAID platform (https://www.gisaid.org/). The complete genomes with high coverage for downstream analysis were selected. All retrieved genomes were aligned and annotated using ON563414.3 as the reference genome by MAFFT ^11^ and VAPID ^12^ tools, respectively. Functional categories were designated using the OPG number previously reported ^13^. The lineages assignment was carried out using the Nextclade web server ^14^.

### Phylogenomic and phylogeographic analyses

We conducted a Maximum Likelihood (ML) phylogeny reconstruction on the genome’s multiple sequence alignment (MSA) and selected the appropriate nucleotide substitution model (GTR+I+G) using IQ-TREE ^15^. A time-scaled phylogenetic tree was built using TreeTime ^16^ to determine the migration dynamics of MPXV and the possible introductions of the virus to Mexico. Additionally, TreeTime was used to infer MPXV introductions to Mexico, with the mugration model, which performs an ancestral character reconstruction of candidate countries of origin as discrete characters. We used Snippy (https://github.com/tseemann/snippy) to detect single-nucleotide variations using the ON563414.3 as a reference genome.

Additionally, to infer the introductions of MPXV to Mexico, PastML was also used ^17^. PastML employed ML marginal posterior probabilities approximation (MPPA) and Felsenstein 1981 substitution model options to reconstruct ancestral character states and their changes along with the trees. As prediction method parameters in PastML, we chose MPPA with standard settings and included character predictions from the joint reconstruction, even if they were not initially selected by the Brier score, using the “forced_joint” option.

### Natural selection analysis

Homologous groups (HGs) were identified using roary ^18^, with BLAST search parameters set to a length coverage of ≥ 90 and amino acid sequence identity of ≥ 90. Subsequently, HGs containing ≥ 10 sequences were selected, and these were aligned with MUSCLE ^19^, specifying 50 iterations. To create a DNA alignment in frame of each HG, the program TRANALING was used ^20^. Then, the presence of natural selection signal (ratio of synonymous/non-synonymous substitutions) was evaluated on the 194 HGs using the FELmethod ^21^, which is available on the Datamonkey server ^22^.

### Ancestral state reconstruction

Ancestral states for the amino acids of the protein encoded by the Cop-A4L gene were reconstructed in Mesquite 3.81 (http://www.mesquiteproject.org), using a parsimony model with mapping of the inferred ancestral states on the ML phylogeny of the whole MPXV genome.

## RESULTS

### MPXV Introductions in Mexico Followed by Local Propagation

On May 28, 2022, the first case of a patient with MPXV was reported in Mexico. However, according to health authorities, it was a patient residing in New York State, USA ^23^. Therefore, to identify the introduction events of MPXV in Mexico, we collected a total of 880 MPXV genome sequences from GISAID on August 2022, for all Mexican MPXV isolates were considered until December 2022 (Supporting information 1). This collection is broad in time, spanning 1970 to 2022, and is also geographically diverse, as MPXV genomes originate from 41 locations (40 countries and the United Kingdom) corresponding to five continents (Africa, Europe, America, Asia, and Oceania). From this collection, 17 genomes were collected from Mexico (Supporting information 1). Using Nextclade ^14^, we performed the lineage designations of the dataset. In the lineages recovered globally, we found that the circulating MPXV population was mainly composed of 24 different lineages, of which the B.1 lineage was the most frequent (52% of the genomes), followed by B.1.6, which corresponded to only 7.32% of the genomes (Supporting information 1). Interestingly, this lineage is only found in isolates from Peru, where they represent 79.26% of the samples.

The phylogenetic reconstruction of the complete genome of MPXV revealed the presence of three main clades (Supporting information 2). Namely, clade I and IIa, which correspond to the Congo Basin lineage and Liberia, respectively. Clade IIb (blue branches) corresponds to cases from West Africa, England, Israel, Singapore, USA (cases reported from 2017 to 2021) and the current outbreak of 2022. We observed that the sequences obtained from Mexico were not confined to a single branch but were scattered throughout the tree (red stars), which could indicate multiple introductions of the virus in the country.

In order to elucidate the introduction of MPXV into Mexico, we used a phylogeographic approach in combination with geographic and temporal information from MPXV genomes and the mugration model implemented in TreeTime, which can be used to describe the migration history of a virus, track its spread, and identify the source of introduction. Initially, with time and the dates of collection of MPXV samples, the time to the most recent common ancestor (tMRCA) of all our included samples was estimated by simple least squares regression, resulting in an inferred date of the MRCA for MPXV of 1962 with a substitution rate of 3.85e^-05^ (Supporting information 3).

The ancestors of the branches containing Mexican isolates were traced back to the United Kingdom (Figure 1: A-C), Portugal (Figure 1: D), and Canada (Figure 1: E), indicating potential sources of introduction of the sequenced MPXV samples into Mexico. Interestingly, MPXV genomes with ancestors from the United Kingdom were distributed across various branches, suggesting multiple introductions.

**Figure 1.**
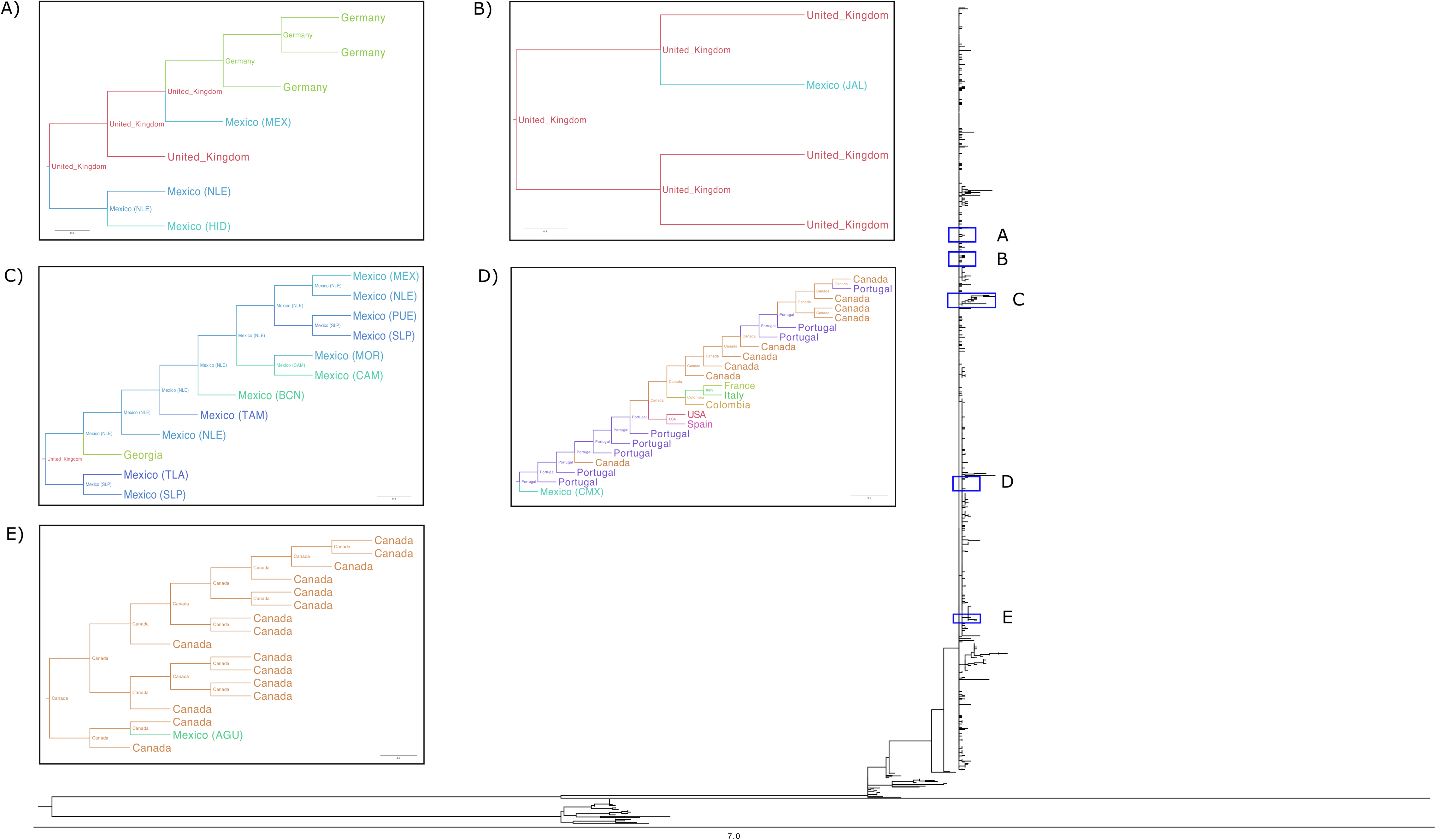
Scaled phylogenetic tree of MPXV introductions to Mexico. 5 clades are observed showing the MPXV introductions in (A and B) Zoom-in on of the clade with introductions from the UK. (C) Zoom-in on the clade with introductions from the UK and with community transmission. (D) Zoom-in on the clade with an ancestor from Portugal. (E) Zoom-in on the clade with ancestors from Canada.

Notably, a distinctive cluster of 11 genomes was identified (Figure 1: C), possibly representing a community transmission event from which the virus spread within Mexico. Remarkably, the sequences within this clade from nine different locations in Mexico, including the State of Mexico (MEX), Nuevo León (NLE), Puebla (PUE), San Luis Potosí (SLP), Morelos (MOR), Campeche (CAM), Baja California Norte (BNC), Tamaulipas (TAM), and Tlaxcala (TLA). Despite being geographically distant, these sequences formed a cohesive cluster within the phylogenetic tree, strongly suggesting widespread transmission across Mexico following their introduction from the United Kingdom.

Our phylogenetic analysis also highlighted independent introductions of MPXV from Portugal and Canada into Mexico City (CMX) and Aguascalientes (AGU), respectively (Figure 1 D and E). The presence of MPXV genomes with ancestors from these two countries in a single branch suggests separate introduction events.

Using TreeTime, the ancestral location of the MPXV sequences of clade IIa was identified in Liberia, whereas for the genomes belonging to clade IIb, which constitute the current outbreak, the ancestral location is inferred to be Nigeria, from where it spread to Germany, Portugal, and the United Kingdom. Additionally, using PastML, we inferred that 11 sequences that formed a cluster in Mexico originated in the United Kingdom, confirming the TreeTime results (Supporting information 4).

### Gene Content Variation and Pangenome Inference

In order to investigate the pangenome of MPXV, an analysis of the coding genome was performed using 880 genomes. The complete coding genome consisted of 158,659 distinct genes grouped into 239 homologous groups (HGs). Among these, 18 HGs were identified as core genes, while 147 HGs belonged to the soft-core genes. Core genes are defined as genes that are present in all isolates, whereas soft-core genes are present in at least 95% of genomes. Additionally, the accessory genome, known as the cloud genome, accounted for 22.59% of the HGs of MPXV (as shown in Supporting information 5). This suggests that the vast majority of MPVX genes belong to either the core genome or the soft-core. Furthermore, the distribution of gene clusters in a bimodal, asymmetrical U-shape pattern (Supporting information 6) indicates that most of the genes in MPXV are either rare or present in nearly all genomes. This observation suggests that MPXV has an open pangenome. Indeed, the total number of genes does not increase significantly as more genomes are examined but never completely stops increasing, which is one of the hallmarks of an open pangenome. To determine the size of the pangenome, we performed 100,000 permutations and obtained an alpha value of 1.0, based on protein coding. These results are further evidence that MPXV exhibits an open pangenome, where the pangenome is either open if α ≤ 1 or closed if α > 1 ^24^. Moreover, considering only the sequences of the current outbreak (clade IIb), the alpha parameter also demonstrates an open pangenome (α = 1.0). The findings indicated that the result of the pangenome analysis was not affected by the sequences from distant clades (Clade I and IIa).

On the other hand, it is important to highlight that some genomes (6.81%) showed a notorious gene variation. In Figure 2, we can clearly see that the genomes of clade I and IIa are grouped together, separated from the genomes of cluster IIb based on their homologous content. In general, we observed a slight trend of the absence of homologs in the right terminal region. However, two clusters of genomes belonging to clade IIb (red rectangles) are also formed (Figure 2). These clusters showed a greater absence of homologs compared to the rest of the isolates, and the fact that some of the genes (homolog clusters) were only present in a few genomes. The first cluster (from the bottom upwards in Figure 2), composed of 41 isolates, did not correlate with isolation place (country) or lineage, which would have potentially explained the gene presence/absence of these isolates. On the other hand, the second cluster is composed of 19 isolates mainly from Belgium (corresponding to 56.25% of the sequences isolated in Belgium). Gene loss in these clusters was observed across all three regions of the MPXV genomes. Moreover, these gene losses and gains occurred in all functional categories.

**Figure 2.**
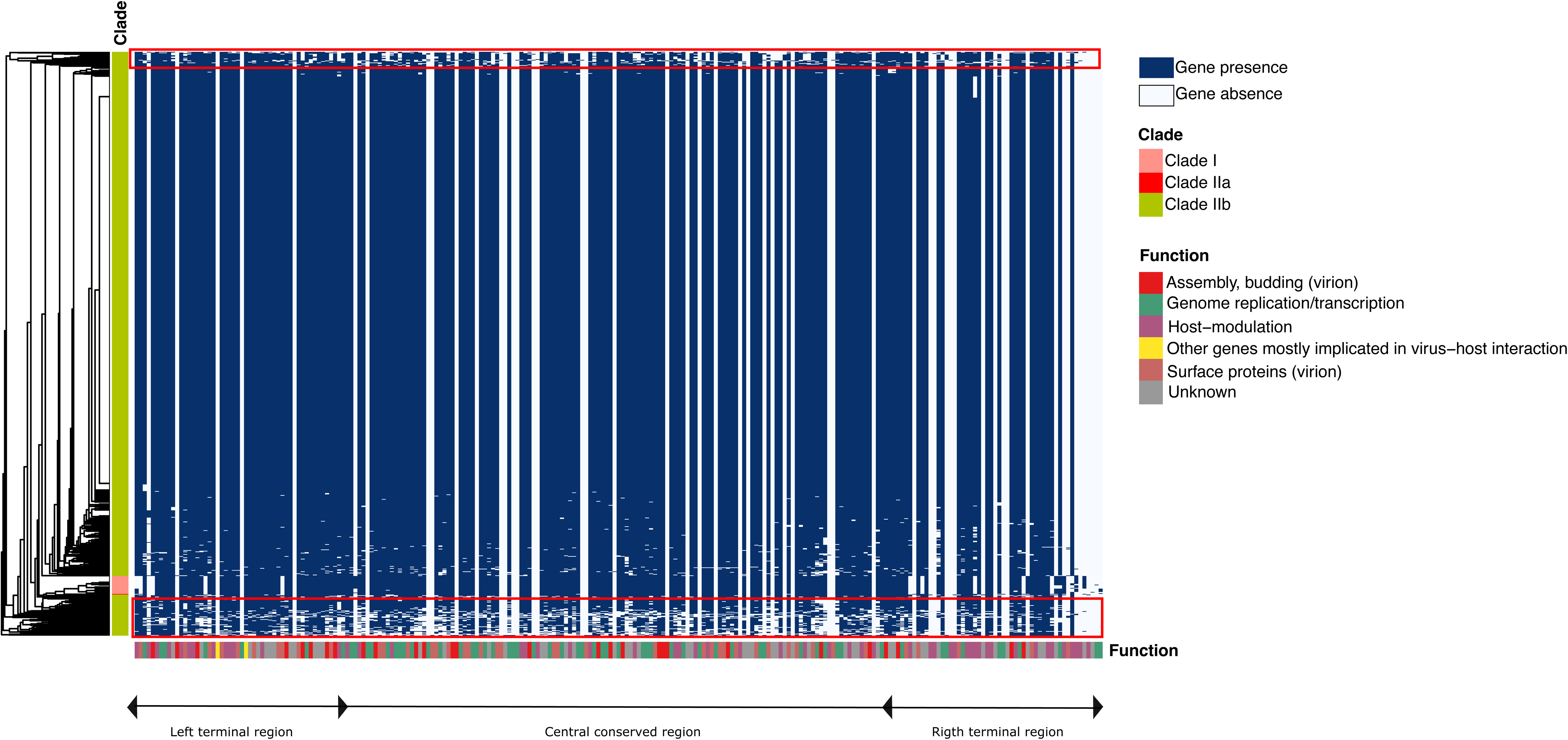
(A) The barplot shows the total number of SNPs for the genes of each functional category. SNPs were identified using GenBank accession ON563414.3 as a reference genome. (B) Boxplot of the number of sites under purifying selection per gene per functional category. Natural selection was evaluated on the 194 HGs using FEL method (see methods). The color coding for each functional category is shown in the figure.

Therefore, the idea of gene turnover at the ends of the MPXV genomes was not clearly observed in our dataset ^8^. However, we can highlight that clusters 1 and 2 (Figure 2; red rectangles) contained groups of homologs coding for different categories and were not present in most clade IIb isolates.

### Evolutionary processes in the Monkeypox virus

Previous reports have shown that the number of SNPs ^25^ in genomes isolated in 2022 increased 6-12-fold compared to those isolated in 2018-2019. Here, we identified SNPs, insertions, and deletions in the 879 MPXV genomes, using the 2018 strain (ON563414.3) as reference. As we expected, sequences of clade I (27 sequences) show more SNPs, insertions, and deletions (Table 1). A total of 5,788 SNPs were observed in the sequences of clade IIb, resulting in seven SNPs per genome, more than one would expect considering previous estimates of 1–2 substitutions per genome per year for Orthopoxviruses ^26^.

Genetic mutations play a crucial role in shaping genomic diversity within natural populations. To gain insights into the selective regimes operating on MPXV, we looked for evidence of diversifying or purifying selection and neutral evolution in the virus using a codon-based phylogenetic framework. The ratio of non-synonymous (dN) to synonymous (dS) substitutions per coding sequence site (dN/dS) was calculated, allowing assessment of the selective pressures acting on specific regions of the MPXV genome and providing valuable information about its adaptive strategies and potential interactions with hosts. Natural selection analyses at each of the genome sites revealed that the sites are highly conserved; out of a total of 194 analyzed groups of homologous coding genes, 277 sites were found to have evolved under purifying selection, distributed across 102 gene groups, particularly the genes responsible for genome replication/transcription and host modulation. Most of the sites that evolved under purifying selection were found within genes located in the core region of the genome and at the 5’ and 3’ ends of the genome. Similarly, the sites that evolve under neutrality were also found mainly within the gene groups responsible for genome replication/transcription and host modulation, with 928 and 887 sites, respectively (Figure 3).

**Figure 3.**
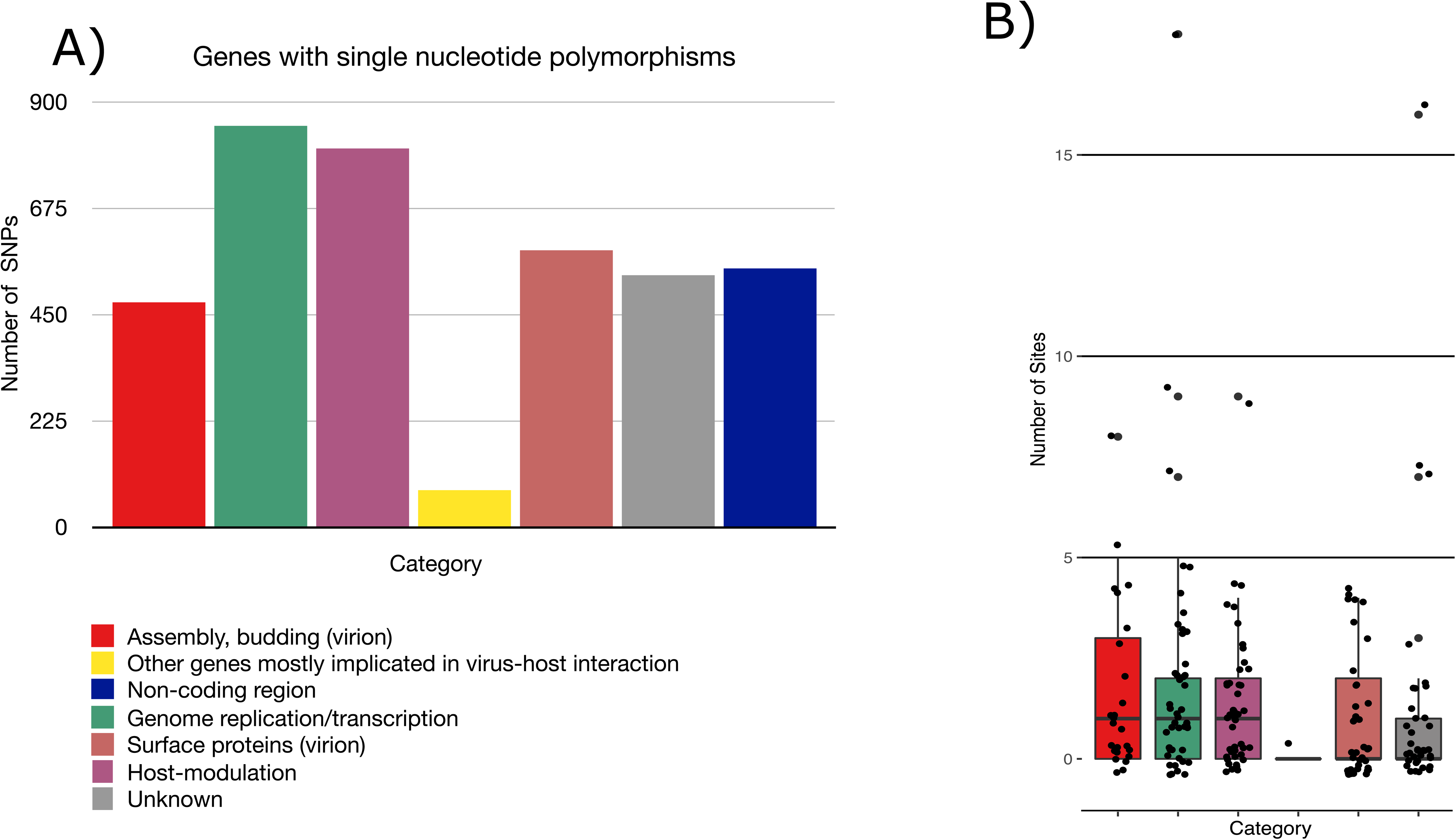
Ancestral state reconstructions using maximum parsimony in Mesquite. Ancestral states for the Cop-A4L gene were mapped onto MPXV whole genome ML-phylogeny. Branches are color-coded to represent different amino acids at position 63 Cop-A4L gene.

Furthermore, we found a non-synonymous mutation throughout the genome (dN/dS > 1); the codon site 63 of the Cop-A4L gene yielded a signal of having evolved under positive selection. To investigate ancestral character states, we traced the amino acid site 63 of the protein encoded by the Cop-A4L gene mapping it on the whole genome phylogeny (see methods). The analysis was aimed at disclosing which amino acid was present at that position in a reconstructed ancestral sequence corresponding to the ancestor of all the genomes included in this study and at internal nodes, where descendants split from the common ancestor. A maximum parsimony reconstruction indicated that the ancestral sequences carried a Valine at position 63 (Figure 4) Furthermore, Valine is conserved through all clade I (green branches), and Alanine is present in the IIa clade (red branches). However, isolates with Threonine at position 63 evolved in the origin of clade IIb, to which the current outbreaks of 2022 and cases from 2017 to 2021 belong (Figure 4; blue branches).

## DISCUSSION

We performed phylogenetic and phylogeographic analyses to understand the introduction of MPXV into Mexico and to study evolutionary processes, such as natural selection of the virus. The first case of MPXV reported in Mexico on May 28, 2022, raised concerns about the origin and spread of the virus in that country. Using a large dataset of 880 GISAID MPXV genome sequences spanning multiple decades and geographic regions, a phylogenetic tree was reconstructed that traced the migration history of the virus. The phylogenetic reconstruction revealed the presence of three major clades within the MPXV genomes. Clades I and IIa were associated with the Congo Basin lineage, and Clade IIb contained cases from West Africa, England, and the current outbreak in 2022, which spread to many countries. This may be because MPXV experienced a stochastic evolutionary event, that led to further diversification of MPXV and contributed to its spread different geographic locations and its possible adaptation to new hosts within these regions ^27^

Our results reveled that the ancestors of the branches containing Mexican isolates could be traced to the United Kingdom, Portugal, and Canada. This suggests that the sequenced MPXV genomes in Mexico originated from these countries, possibly through travel-associated transmission, as has been observed for other viruses such as SARS-CoV-2, where multiple introductions occur, and only a few viruses manage to establish local transmission chains throughout the country ^28,29^.

Regarding genome evolution, recent evidence suggests that MPXV exhibits gene turnover at the end of its genome ^8^. Here, we found that gene loss and gain in MPXV genomes were scattered throughout the genome. However, we identified a few unique homologous groups (HGs) in clades I and IIa genomes, leading to clustering based on the HG content. Notably, only 6.81% of the genomes from the 2022 outbreak (clade IIb) exhibited a noticeable loss of HGs along their genomes. However, we found no correlation between the isolation site (country) or lineage and loss of HGs in these genomes. This suggests the possibility that these results may be attributed to sequencing artifacts or other factors rather than to inherent viral biology.

On the other hand, we found that MPXV has an open pangenome, which means that as the number of genomes increases, new genes are likely to be discovered. These observations suggest that gene turnover may play a crucial role in shaping the genomic diversity of MPXV. In addition, it has reported that inverted repeats in the MPXV genome also could be hotspots of genetic variability in MPXV genomes ^25^.

Furthermore, we noticed a higher number of SNPs than in previous reports, taking into account the rate of 1-2 genetic substitutions per year for Orthopoxviruses ^26^.

However, these variations are unlikely to affect the diversity of the viral genome, and many polymorphisms are subject to purifying selection. Overall, the results have shown that sites within the genome evolve largely under purifying selection. This is consistent with the findings of Yong Zhan *et al*. (2023), who found that more than 60% of genes evolve under negative selection in the genome of MPXV. Negative selection is responsible for eliminating these deleterious mutations in genes and thus maintaining functional stability. Consequently, it is common to find genes with relevant functions in viruses under this type of selection, such as DNA-binding proteins or DNA-binding proteins, telomeres, virion core proteins, and transcription factors ^30^.

In contrast to Yong Zhan et al. (2023), who reported 10 genes under positive selection that were involved in host association, in our work, we found one site under selection within the Cop-A4L gene. The site V63A/T evolved under diversifying (positive) selection within the gene Cop-A4L. The gene Cop-A4L produces the 39-kDa protein, a highly antigenic protein of the viral core, implicated in the progression of the immature virion to the infectious intracellular mature virion ^31,32^ through interaction with p4a (which is encoded by the A10L gene). This interaction is maintained with the processed 4a from that arises during virion maturation ^33^ (observed in pulse chase and immunoprecipitation experiments). Based on experiments in Vaccinia Virus 39-kDa gene (Western Reserve strain), the 39-kDa protein is located within the last 103 amino acids at the C-terminal. This region induces strong humoral immune responses in humans and animals ^34^. In addition, ancestral state reconstruction deduced that the ancestral state at site 63 corresponds to the amino acid valine, which is only present in isolates of clade I. On the other hand, isolates from the current outbreak (clade IIb) and from clade IIa, encoded for threonine and alanine at site 63, respectively. Alanine and Valine are nonpolar aliphatic amino acids, whereas Threonine is an uncharged polar amino acid. Because the 39-kDa protein (encoded by the Cop-A4L) is disordered, it is difficult to identify whether this mutation could directly affect the structure of the protein and influence the host immune response and/or interaction with p4a during virion maturation. Future analyses are needed in order to understand the impact of T63 on the spread of the current outbreak.

In summary, this study sheds light on MPXV evolution and reveals its multiple introductions into Mexico and the dynamics of its genetic makeup. Our findings contribute to our understanding of the genetic characteristics of the evolution of MPXV.

## Supporting figures legends

**Supporting information 1.** Genomes used.

**Supporting information 2.** Maximum likelihood phylogenetic tree using 880 MPXV genomes. The phylogeny is annotated by Clade I, Clade IIa, and Clade IIb, which are indicated by yellow, green, and blue branches, respectively. The lineage of each isolate is highlighted in different colors (ribbon color). Mexican isolates are denoted with red stars. Abbreviations: Aguascalientes (AGU), Baja California Norte (BCN), Campeche (CAM), Mexico City (CMX), Hidalgo (HID), Jalisco (JAL), State of Mexico (MEX), Morelos (MOR), Nuevo León (NLE), Puebla (PUE), San Luis Potosí (SLP), Tamaulipas (TAM), Tlaxcala (TLA).

**Supporting information 3.** Root-to-tip regression analyses based on the ML trees obtained for MPXV

**Supporting information 4.** Ancestral reconstruction of MPXV. The figure shows the compressed visualization produced by PastML using marginal posterior probability approximation (MPPA) with an F81-like model. Different colors correspond to different geographical regions. Numbers inside (or next to) the circles indicate the number of strains assigned to the specific node.

**Supporting information 5.** Table of counts of shared genes (core genome) and total genes (pangenome).

**Supporting information 6.** Pangenome plot of the global gene repertoire of Monkeypox genome.

## ACKNOWLEDGMENT

Gamaliel thanks Paola Rojas-Estévez for her useful comments on the manuscript.

## CONFLICT OF INTEREST

The authors declare no conflict of interest.

## DATA AVAILABILITY STATEMENT

The original contributions presented in the study are included in the article/Supporting Information; further inquiries can be directed to the corresponding authors.

---

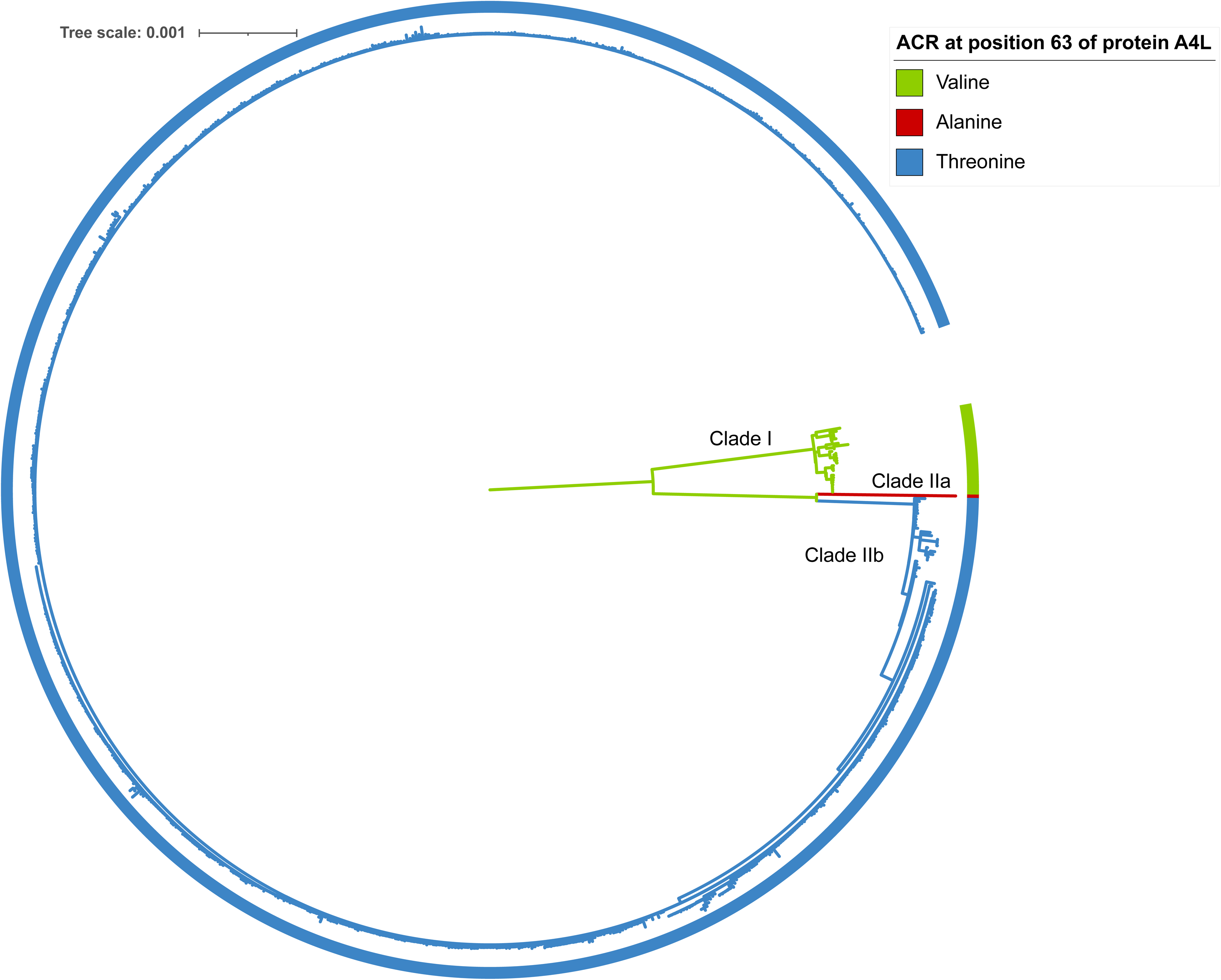

## Notes

### Competing Interest Statement

The authors have declared no competing interest.

### Funding Statement

This study did not receive any funding

### Author Declarations

The study used ONLY openly available human samples.

